# Selective 10-Hz photic driving deficit shared between Alzheimer’s and frontotemporal dementia

**DOI:** 10.64898/2026.07.23.26358714

**Authors:** Pedro Velez-Pardo, Ricardo Montoya Monsalve, Daniel Vasquez, Sofía Campuzano Cortina, Eduardo Montoya Guevara

## Abstract

**Background:** Photic stimulation is a routine clinical probe of posterior alpha circuits, yet its signature in frontotemporal dementia (FTD) and subject-level comparison with Alzheimer’s disease (AD) remain uncharacterized.

**Methods:** We analyzed two OpenNeuro datasets sharing 88 participants (86 retained: 35 AD, 29 controls [CN], 22 FTD): resting eyes-closed and photic stimulation at 5, 10, 15, 20 Hz. After ICA preprocessing we computed a photic driving index (DI), resting spectral features, and weighted phase-lag connectivity (wPLI). Linear mixed-effects models tested group effects; leave-one-subject-out cross-validation with five classifiers quantified diagnostic utility.

**Results:** DI at 10 Hz was reduced ∼50% in AD (*β* = −6.96, *p* = 0.004) and FTD (*β* = −6.74, *p* = 0.012), robust under log, winsorized, and rank-based sensitivity analyses. Resting analyses showed 1.5 Hz IAF slowing in AD and posterior theta/alpha ratio elevation up to 6-fold (AD) and 3.3-fold (FTD). wPLI showed no group differences, but equivalence was not established (TOST *p* ≥ 0.16). DI split-half reliability at 10 Hz was near null (ICC = 0.06). Balanced accuracy was 0.577 for resting versus 0.543 combined (McNemar *p* = 0.85).

**Conclusion:** A selective alpha-band amplitude deficit is shared between AD and FTD but adds no diagnostic value over resting EEG. Distinguishing reduced cortical gain from preserved synchrony requires longer recordings powered for equivalence testing.

## 1 Introduction

### 1.1 Alzheimer’s disease

Alzheimer’s disease (AD) is the leading cause of dementia worldwide, accounting for 60-80% of all recognized cases and affecting more than 50 million people globally[1–3]. Its prevalence approximately doubles every five years after age 65, and projections suggest these figures will double or triple by 2050[3–5]. Clinically, AD follows a prolonged preclinical course spanning years to decades before manifesting as progressive memory-led cognitive decline that later compromises behavior and daily functioning[4, 6–8].

Its defining histopathological hallmarks are extracellular *β*-amyloid plaques and neurofibrillary tangles of hyperphosphorylated tau, driving widespread synaptic and neuronal loss with reactive glial changes[1, 9]. Advanced age and the APOE*ε*4 allele represent the principal risk factors[2, 4]. Contemporary diagnosis combines clinical history, neuropsychological testing, and cerebrospinal fluid or PET biomarkers for A*β* and tau[2, 10]. Approved treatments (cholinesterase inhibitors and memantine) offer only symptomatic relief; monoclonal antibodies targeting amyloid represent a promising disease-modifying approach with logistical challenges[3, 4].

### 1.2 Frontotemporal dementia

Frontotemporal dementia (FTD) encompasses a heterogeneous group of neurodegenerative disorders selectively affecting the frontal and anterior temporal lobes, producing prominent alterations in behavior, personality, executive function, and language[11, 12]. It typically manifests in the fifth or sixth decade of life, though cases span from age 30 to over 90[11, 13], and recent meta-analyses have refined its incidence and prevalence estimates upward[12, 14].

FTD comprises several core syndromes: the behavioral variant (bvFTD), characterized by disinhibition, apathy, and severe social conduct impairment[11, 15]; the primary progressive aphasias including semantic (svPPA) and nonfluent / agrammatic (nfvPPA) variants[11, 12]; and the right temporal variant with prosopagnosia and right anterior temporal atrophy[16]. FTD overlaps clinically and pathologically with motor neuron disease and atypical parkinsonian syndromes[11, 12, 17]. Diagnosis relies on clinical assessment, neuropsychological testing, MRI atrophy patterns, and fluid biomarkers including CSF tau/p-tau and blood neurofilament light chain[15, 17–19]. Despite these tools, FTD remains underdiagnosed and is frequently misattributed to primary psychiatric illness or AD.

### 1.3 Quantitative EEG and photic driving in dementia

Quantitative EEG (qEEG) has long been considered a candidate biomarker for AD, with classical findings of posterior alpha slowing and theta/alpha ratio elevation[20–22]. EEG abnormalities in FTD have been historically described as relatively preserved, a generalization recently challenged by modern quantitative analyses revealing distinct spectral and connectivity signatures in both conditions[23, 24].

Intermittent photic stimulation (IPS) is a routine clinical EEG procedure providing an active probe of posterior alpha circuitry[25]. By presenting periodic visual flickers, it generates a steady-state visual evoked potential (SSVEP) whose amplitude and harmonic structure reflect the integrity of thalamocortical alpha generators[26]. In AD, quantitative studies consistently report reduced alpha power at rest and decreased photic driving at stimulus frequencies (notably 10 and 15 Hz)[27–30]. A blunting of coherence reactivity—the shift from rest to stimulation over posterior regions—reflects failure of normal stimulus-induced cortical activation[31, 32]. AD patients also show selectively reduced harmonic responses and altered multiscale entropy during IPS[33, 34]. Large memory-clinic cohorts confirm that AD patients more often show EEG abnormalities than FTD patients under routine visual inspection[35].

### 1.4 Gap and contribution

Direct IPS investigations in FTD remain scarce, and contemporary machine-learning classifiers for AD-versus-FTD distinction have relied almost exclusively on resting-state EEG[23, 24, 36, 37]. The recent release of two complementary OpenNeuro datasets sharing 88 participants—one in resting-state eyes-closed[38] and another with intermittent photic stimulation at 5, 10, 15, and 20 Hz[39]—opens an unprecedented methodological pathway for direct comparison. To our knowledge, only one prior study has analyzed both jointly[40]; however, it treated the photic dataset as a generic eyes-open condition without extracting SSVEP-specific features. Recent advances in 40-Hz gamma entrainment further support the translational relevance of photic stimulation as both a diagnostic probe and a therapeutic intervention in AD[41–45].

We present a hypothesis-driven secondary analysis with two aims. First, we characterize the photic driving response across groups using linear mixed-effects models that account for repeated measures across stimulation frequencies. Second, we quantify the incremental classification value of photic features over resting features alone using leave-one-subject-out (LOSO) cross-validation with bootstrap confidence intervals[46], with subject-level evaluation designed to avoid the overoptimism documented in prior epoch-level validations.

## 2 Methods

### 2.1 Study design and rationale

This is a hypothesis-driven secondary analysis of two complementary OpenNeuro datasets sharing an identical cohort of 88 clinically characterized participants. The within-cohort cross-dataset design eliminates between-subject variability confounding most published resting-versus-task EEG comparisons. Subject-level (rather than epoch-level) cross-validation was specified a priori as the primary classification framework to avoid over-optimistic accuracy estimates documented in prior machine-learning analyses of these datasets[23, 40, 46].

### 2.2 Datasets and participants

We used `ds004504` (resting-state eyes-closed, mean duration ∼ 13 minutes)[38] and `ds006036` (intermittent photic stimulation at 5, 10, 15, 20 Hz with alternating eye states, mean ∼ 5 minutes)[39], sharing 88 identical participants (of whom 86 completed the analytic pipeline). Both datasets were acquired at AHEPA University Hospital, Thessaloniki, with a 19-channel scalp montage (Nihon Kohden EEG 2100; international 10–20 system; A1–A2 linked-mastoid reference; 500 Hz sampling; analog band-pass 0.5–70 Hz). Photic blocks are marked as `PHOTO XHz` events with individual flash onsets as `Photo/HV mark` events. The original protocol was approved by AHEPA Scientific and Ethics Committee (protocol 142/12-04-2023) and data are distributed under CC0 license. Diagnostic groups were defined per clinical criteria (NINCDS-ADRDA for AD, International Behavioural Variant FTD Criteria Consortium consensus for FTD, MMSE = 30 for CN). Biomarker confirmation was not available in source data; we treat this as a limitation. Demographic characteristics are summarized in Table 1.

**Table 1:**
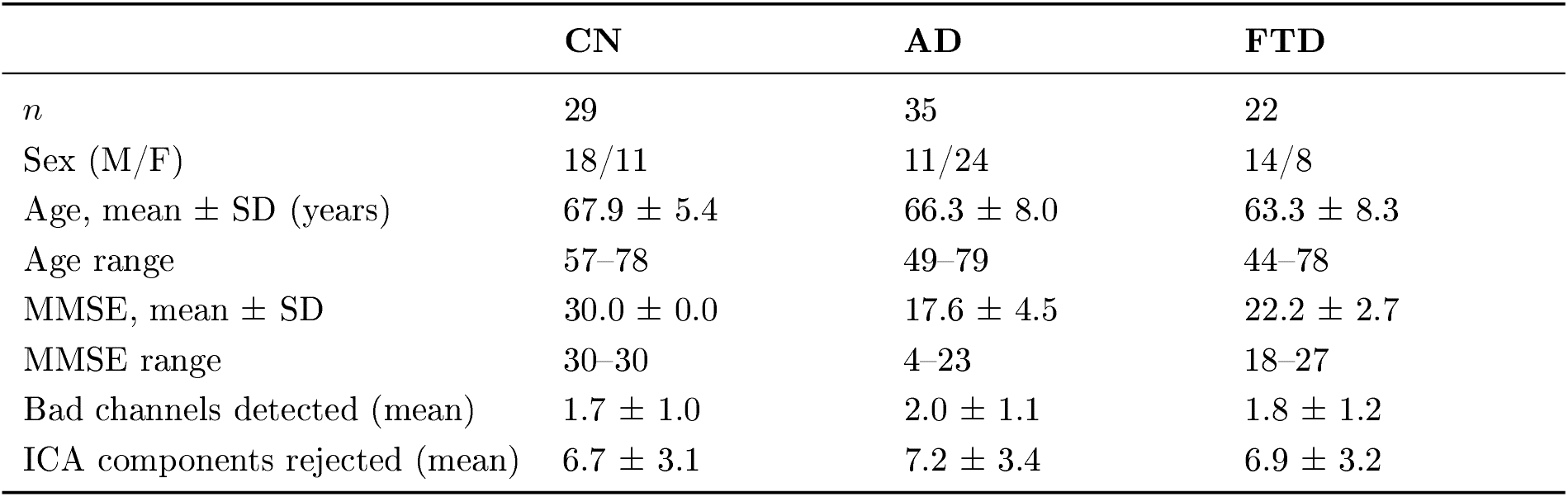
Sample demographics by diagnostic group. Values correspond to the 86 subjects retained in the analytic pipeline (of 88 available in the source datasets). Two subjects were excluded for insufficient recording duration.

|  | CN | AD | FTD |
| --- | --- | --- | --- |
| *n* | 29 | 35 | 22 |
| Sex (M/F) | 18/11 | 11/24 | 14/8 |
| Age, mean ± SD (years) | 67.9 ± 5.4 | 66.3 ± 8.0 | 63.3 ± 8.3 |
| Age range | 57–78 | 49–79 | 44–78 |
| MMSE, mean ± SD | 30.0 ± 0.0 | 17.6 ± 4.5 | 22.2 ± 2.7 |
| MMSE range | 30–30 | 4–23 | 18–27 |
| Bad channels detected (mean) | 1.7 ± 1.0 | 2.0 ± 1.1 | 1.8 ± 1.2 |
| ICA components rejected (mean) | 6.7 ± 3.1 | 7.2 ± 3.4 | 6.9 ± 3.2 |

### 2.3 Preprocessing pipeline

All preprocessing was implemented in MNE-Python 1.12.1[47] from raw . `set` files with fully deterministic settings (random seeds fixed at 42):

#### (i) Filtering

A zero-phase FIR band-pass filter (MNE default, Hamming-windowed, transition bandwidth 0.5 Hz at the 1 Hz cutoff and 5 Hz at the 45 Hz cutoff, filter length automatically determined) was applied with cutoffs at 1 and 45 Hz, followed by a 50 Hz notch. The 45 Hz low-pass renders the 50 Hz notch strictly redundant for the mains fundamental but was retained as protection against harmonic leakage. The 1 Hz high-pass mildly attenuates the delta band, which was considered acceptable given that no spectral analyses in this study target sub-2 Hz activity and that a higher high-pass improves ICA decomposition of cortical signals in the 5–45 Hz range.

#### (ii) Channel renaming and montage assignment

Legacy 10–20 electrodes (T3, T4, T5, T6) were renamed to modern equivalents (T7, T8, P7, P8) enabling application of the MNE `standard_1020` montage.

#### (iii) Bad channel detection

Channels with RMS z-score *>* 3.5 MAD from median were flagged; spline interpolation was applied when ≤ 4 channels were flagged. Subjects with more bad channels were excluded.

#### (iv) Independent component analysis

An extended Infomax ICA[48] with 15 components was fitted (max 200 iterations, seed 42). Ocular components were identified via z-scored correlation of component time courses with Fp1/Fp2 channels (z-score threshold 2.5, MNE default in `find_bads_eog`),and muscle components via topographic spectral signature (z-score threshold 0.5 in `find_bads_muscle`). Retaining approximately 8 of 15 fitted components after rejection preserves the majority of the cortical rank in the interpolated 19-channel montage; this is a moderate rejection rate for clinical EEG shown to protect against removal of neural signal in comparable pipelines[47].

#### (v) Protective rule for stimulation-locked components

Because SSVEPs generate spectrally narrow components that can be misclassified as periodic artifacts, we implemented an explicit rule: candidate components whose dominant power lay within ±1.5 Hz of any stimulation frequency (5, 10, 15, or 20 Hz) were removed from the rejection list and retained. This preserved approximately 0.8 components per subject and is a methodological precaution not implemented in the published descriptor pipeline[27].

### 2.4 Segmentation, epoching, and reliability estimation

Photic block boundaries were identified computationally: a block ended at the first `Photo/HV mark` interval *>* 3× the expected inter-pulse interval. Primary analyses used eyes-closed segments within each photic block (∼ 5 s per frequency per subject), reflecting the standard clinical paradigm under which the driving response is most reliably evoked[27, 29]. Two-second epochs were rejected if peak-to-peak amplitude exceeded 150 *µ*V or if `eye movement` events occurred within the window.

Given the short duration of eyes-closed segments per photic frequency, yielding on average 1–3 clean 2-s epochs per subject-frequency, we conducted a split-half reliability analysis on a representative subsample of 18 subjects (6 per group). For each subject and stimulation frequency, clean epochs were divided into first and second halves, DI was computed on each half, and mean values were correlated across subjects. The Pearson correlation was Spearman-Brown corrected: ICC_SB_ = 2*r/*(1 + *r*). The full distribution of clean-epoch counts is reported in Supplementary Table S1.

### 2.5 Feature extraction

#### Spectral features

Power spectral density (PSD) was estimated via Welch’s method[49] with a 2-s Hanning window (0.5 Hz resolution) . Absolute and relative powers in five bands (delta 1–4, theta 4–8, alpha 8–13, beta 13–30, gamma 30–45 Hz), theta/alpha ratio (TAR), and slow/fast ratio ((delta+theta)/(alpha+beta)) were computed per region of interest (ROI) .

#### Individual alpha frequency (IAF)

IAF was defined as the spectral peak in 7–13 Hz, *estimated on the resting-state recording* (*ds004504*) rather than on the photic recording. This pre-specified choice avoids circular contamination by the SSVEP-driven power at 10 Hz, which coincides with the endogenous alpha band and pulls IAF estimates artefactually toward the stimulation frequency in participants with preserved photic driving.

#### Theta/alpha ratio: dual computation

Because the alpha denominator of the TAR is inflated by the SSVEP at 10 Hz in participants with preserved driving, we computed the TAR both on the resting recording (*resting TAR*, uncontaminated) and on the photic 10 Hz block (*photic TAR*) . Reporting both allows separating the pure posterior-slowing component from the driving-linked component.

#### Photic driving index (DI)

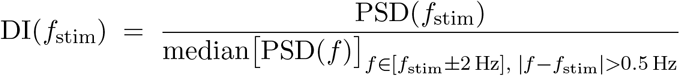

This normalizes the photic response by each subject’s local background spectrum. Harmonic DIs were computed at 2× and 3× the stimulation frequency.

#### Functional connectivity

Weighted phase-lag index (wPLI)[50] in [9, 11] Hz between channel pairs within and between three ROIs: posterior (O1, O2, P3, P4, Pz, P7, P8), central (C3, C4, Cz), and frontal (Fp1, Fp2, F3, F4, F7, F8, Fz). wPLI was preferred over coherence because it suppresses volume-conduction contributions.

### 2.6 Statistical analysis

Analyses were performed in R 4.3 (lme4 1.1.x, lmerTest 3.1.x, emmeans 1.10.x) with Python 3.10 (statsmodels 0.14) for cross-validation. Effect sizes were partial *η*^2^ (with 95% CIs)[51] and Hedges’ *g*.

#### Primary mixed-effects model

For the photic driving index:

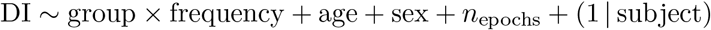

Random intercept was chosen for parsimony given *n*_subject_ = 86; a sensitivity with random slopes yielded equivalent fixed effects with wider CIs. Satterthwaite degrees of freedom were used.

#### Secondary OLS analyses

For variables measured once per subject (TAR, IAF, ROI-pair wPLI), OLS with age, sex, and *n*_epochs_ as covariates. Post-hoc Tukey HSD via emmeans.

#### Equivalence testing

For the AD-versus-FTD contrast where absence of significance is not evidence of equivalence, we conducted two one-sided tests (TOST)[52] with an equivalence margin of ±4 DI units (approximately half the CN–AD difference) .

#### Multiple comparisons

Benjamini–Hochberg FDR[53] across the four stimulation frequencies within each outcome. Outcomes (DI, TAR, IAF, wPLI) were not pooled because each addresses distinct hypotheses.

#### Handling of MMSE

MMSE was used only descriptively and in a within-AD severity analysis; because MMSE separates CN from AD by definition (ceiling effect in CN), including it as a covariate in group-difference models would produce mathematical collinearity.

### 2.7 Classification framework

Leave-one-subject-out (LOSO) cross-validation eliminates the leakage arising from epoch-level splitting[46]. Five classifiers spanning interpretable to flexible model classes were evaluated: logistic regression with elasticnet penalty (*ℓ*_1_ ratio 0.5), SVM with RBF kernel (*C* = 1, *γ* = scale), random forest (300 trees), gradient boosting (200 estimators), and XGBoost[54]. Three feature sets: resting-only (42 features), photic-only (186), combined (228). Median imputation and standard scaling were fitted on training data only within each fold.

For each combination we report balanced accuracy (primary), macro-F1, and one-vs-rest AUC. SVM-RBF was a priori declared the primary clinical-grade model. XGBoost yielded a one-vs-rest AUC of 0.33, *below* the chance-level AUC of 0.5[55], indicating systematically miscalibrated (effectively inverted) class probabilities despite a valid balanced-accuracy point estimate. This calibration failure is a known interaction between multiclass label encoding and class-weight balancing in histogram-based boosters; XGBoost is therefore reported only as a sensitivity model.

Bootstrap 95% CIs were estimated by resampling subject-level predictions with replacement 10,000 times. A permutation test (500 label permutations) tested the null of chance-level balanced accuracy for the SVM-RBF combined model. The pre-specified McNemar exact test on per-subject correctness vectors directly compared resting-only vs combined models.

### 2.8 Sensitivity analyses

Eight pre-specified sensitivity analyses examined robustness: (i) exclusion of AD *<* 55 years; (ii) exclusion of FTD *<* 55 years; (iii) exclusion of AD with MMSE *<* 10; (iv) male-only stratum; (v) female-only stratum; (vi) ≤ 3 interpolated bad channels; (vii) ≤ 10 ICA components rejected; (viii) comparison between the ICA-based primary pipeline and a minimal pipeline (Supplementary Table S2). Following external methodological review, we added (ix) three post-hoc distributional robustness checks for the primary DI @ 10 Hz mixed-effects model: log-transformation of the outcome (log(1 + DI)), 5/95% winsorization per stimulation frequency, and a rank-based Kruskal–Wallis test with Bonferroni-corrected post-hoc contrasts (Supplementary Table S3). We also added (x) a TOST equivalence test on the fronto-posterior wPLI contrast using a ±0.5 SD margin, matching the equivalence margin used for the AD-versus-FTD DI comparison.

### 2.9 Open science and reproducibility

The complete analysis pipeline (protocol documents, preprocessing/statistical scripts, LOSO framework, feature matrices) was deposited in a private GitHub repository at execution time and timestamped via Git commits; the repository will be made public upon acceptance with tag `v1.0-submission` [56]. We did not use a formal pre-registration platform; the audit trail of timestamped commits and intermediate result files in the GitHub repository serves as the equivalent transparency artifact. Software versions: MNE-Python 1.12.1, scikit-learn 1.7.2, scipy 1.13, statsmodels 0.14, R 4.3.

## 3 Results

### 3.1 Sample characteristics and data quality

Of 88 participants in the source datasets, 86 (97.7%) successfully completed the ICA-based pre-processing pipeline and were retained for primary neurophysiological analyses (35 AD, 29 CN, 22 FTD; Table 1). Two subjects (one AD, one FTD) were excluded for recording duration *<* 60 s in the resting condition. Cross-dataset matching was performed exclusively by the BIDS participant identifier (`sub-XXX`), which is preserved between `ds004504` and `ds006036`; identifier equality therefore guarantees within-subject matching without recourse to demographic or acquisition-date fields. After cross-matching photic and resting datasets, 83 subjects (34 AD, 28 CN, 21 FTD) had complete feature vectors for combined LOSO classification.

Groups did not differ in age (Kruskal-Wallis *H* = 2.62, *p* = 0.27) but differed in sex distribution (*χ*^2^ = 6.18, *p* = 0.046): AD was enriched for females (24/35, 69%) compared with CN (11/29, 38%) and FTD (8/22, 36%). All inferential models were adjusted for sex. Data quality was uniformly high: mean 1.84 ± 1.10 bad channels per recording (range 0–4); ICA rejected mean 6.95 ± 3.24 of 15 fitted components (Kruskal–Wallis *p* = 0.41 across groups). The protective rule preserved on average 0.8 components per subject.

### 3.2 Selective alpha-band photic driving deficit at 10 Hz

Descriptive statistics of the DI in posterior channels revealed marked group separation specifically at 10 Hz (Figure 1A). CN showed a mean DI of 13.24 ± 16.05, nearly double that of AD (6.49 ± 6.33) and FTD (6.62 ± 7.63); reductions relative to CN were 51% in AD and 50% in FTD. Distributions largely overlapped at 5 Hz (CN 3.90 ± 2.92; AD 4.02 ± 4.16; FTD 3.95 ± 3.86), 15 Hz (CN 5.45 ± 4.29; AD 7.06 ± 6.02; FTD 7.68 ± 12.80), and 20 Hz (CN 7.73 ± 7.45; AD 6.73 ± 7.94; FTD 6.38 ± 6.51).

**Figure 1:**
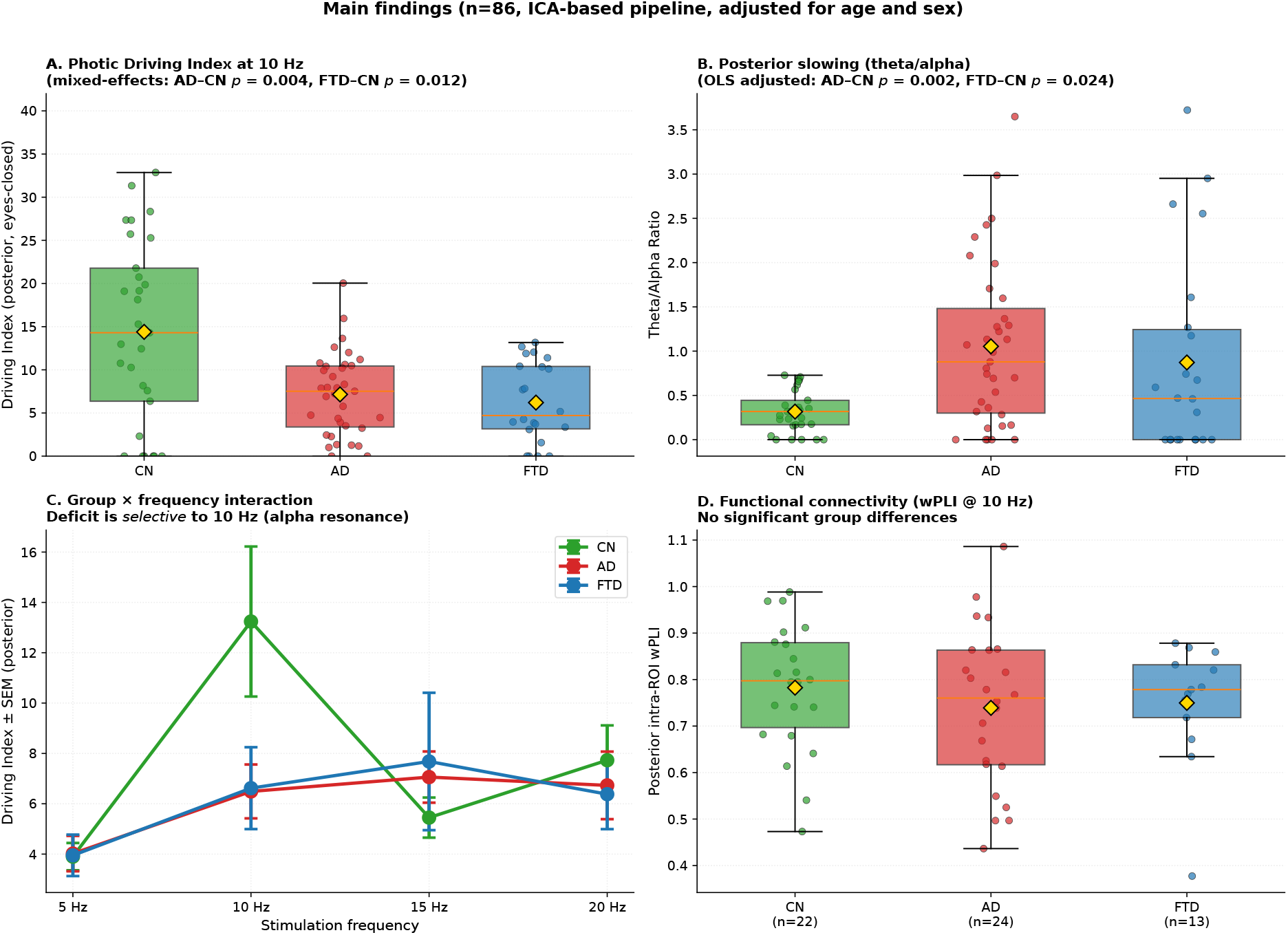
Main findings. (A) Photic driving index at 10 Hz, reduced approximately 50% in both AD and FTD relative to CN. (B) Posterior theta/alpha ratio during 10 Hz stimulation, elevated in both dementia groups. (C) Driving index by stimulation frequency, showing the deficit is selective to 10 Hz (alpha resonance) with no differences at 5, 15, or 20 Hz. (D) Weighted phase-lag index at 10 Hz in posterior intra-ROI connectivity, with no group differences detected; formal equivalence, however, was not established (Section 3.5).

The linear mixed-effects model confirmed selective 10 Hz interactions (Table 2). Main effect of frequency was highly significant (*F* (3, 240.8) = 9.65, *p <* 0.001), driven by elevated DI at 10 Hz vs 5 Hz in CN (*β* = +9.32, *p <* 0.001). Interaction terms at 10 Hz reached significance for both AD (*β* = −6.96, 95% CI [−11.64, −2.27], *p* = 0.004) and FTD (*β* = −6.74, [−11.99, −1.49], *p* = 0.012), indicating attenuation of natural alpha-band resonance in both dementia groups.

**Table 2:** Statistical models for primary outcomes. Adjusted for age and sex; mixed model also adjusted for number of clean epochs. Significance markers: ^*^*p <* 0.05, ^**^*p <* 0.01.

| Outcome | Effect | $\beta$ | SE | 95% CI | *p* |
| --- | --- | --- | --- | --- | --- |
| *Theta/Alpha ratio at PHOTO 10 Hz (OLS adjusted)* |  |  |  |  |  |
|  | AD vs CN | +0.78 | 0.24 | [0.31, 1.26] | 0.0015** |
|  | FTD vs CN | +0.59 | 0.26 | [0.08, 1.10] | 0.024* |
| *Driving Index, mixed-effects model (group × frequency interaction)* |  |  |  |  |  |
| | AD × 10 Hz | −6.96 | 2.38 | [−11.64, −2.27] | 0.0038** |
| | FTD × 10 Hz | −6.74 | 2.66 | [−11.99, −1.49] | 0.012* |
| *IAF on resting recording (OLS adjusted)* |  |  |  |  |  |
|  | AD vs CN | −1.54 | 0.39 | [−2.31, −0.77] | < 0.001** |
|  | FTD vs CN | −0.69 | 0.43 | [−1.54, +0.16] | 0.110 |
| *TOST equivalence AD vs FTD at DI 10 Hz (margin $\pm 4$)* |  |  |  |  |  |
|  | AD − FTD | −0.13 | — | [−4.07, +3.81] | 0.027* (equivalence) |

Interactions at 5, 15, and 20 Hz were all non-significant (*p >* 0.39), supporting spectral selectivity (Figure 1C).

The AD-versus-FTD contrast at 10 Hz was not significant (Hedges’ *g* = −0.02, 95% CI [−4.07, +3.81]). Because absence of a significant difference is not evidence of equivalence, we conducted a TOST equivalence procedure with an equivalence margin of ±4 DI units. TOST rejected the null of a difference exceeding the margin (upper *t* = −2.12, *p* = 0.020; lower *t* = +1.99, *p* = 0.027), providing formal statistical support for equivalence within this pre-specified margin.

Split-half reliability analysis revealed that DI at 10 Hz has essentially null within-subject reliability (Pearson *r* = 0.03, ICC_SB_ = 0.06, *p* = 0.92; *n* = 11 subjects with ≥ 2 epochs per half). Given the small subsample, the reliability estimate itself is imprecise: the Fisher-*z* 95% confidence interval spans [−0.56, +0.64], meaning the point value should be read as “low reliability, magnitude uncertain” rather than as a precise null. The qualitative conclusion of low individual-level reliability is nevertheless robust to this uncertainty because the confidence interval excludes reliability values compatible with subject-level biomarker use (≥ 0.75). Reliability at 5 Hz was moderate (ICC = 0.66, *p* = 0.15). Subjects averaged 1–3 clean 2-s epochs per stimulation frequency (Figure 4A). This population-versus-individual dissociation—a genuine group-level effect coexisting with unreliable individual estimates—fundamentally constrains the metric’s utility as a subject-level biomarker and mechanistically foreshadows the null classification results (Section 3.7).

#### Sensitivity to distributional assumptions

Because the DI has a right-skewed distribution with occasional extreme values, we refit the primary mixed-effects model under three robust specifications: log-transformation of the outcome (log(1 + DI)), 5/95% winsorization per frequency, and a rank-based Kruskal-Wallis test. The marginal group contrast at 10 Hz remained significant in all three: under log-transformation *β*_AD_ = −0.53 (*p* = 0.010) and *β*_FTD_ = −0.59 (*p* = 0.009); under winsorization *β*_AD_ = −3.40 (*p* = 0.028) and *β*_FTD_ = −3.46 (*p* = 0.041); the rank-based Kruskal–Wallis was significant globally (*H* = 7.44, *p* = 0.024) with marginal Bonferroni-corrected differences for CN vs AD (*p* = 0.067) and CN vs FTD (*p* = 0.055), and no evidence of an AD vs FTD difference (*p* = 1.00). Full sensitivity results are reported in Supplementary Table S3. The primary finding therefore does not depend on the influence of a few extreme values.

### 3.3 Theta/alpha ratio elevation in dementia

Posterior TAR during 10 Hz photic stimulation was elevated in both AD (mean 1.04 ± 1.04, 3.7-fold vs CN) and FTD (0.84 ± 0.99, 3.0-fold vs CN) relative to CN (0.28 ± 0.26; Figure 1B). Adjusted OLS: AD-CN *β* = +0.78, 95% CI [0.31, 1.26], *p* = 0.0015; FTD-CN *β* = +0.59, [0.08, 1.10], *p* = 0.024 (Table 2). Age and sex covariates were non-significant. The effect was robust across eight pre-specified sensitivity analyses (AD-CN *p* range 0.001-0.034).

To address the concern that the SSVEP might inflate the alpha denominator and spuriously exaggerate the effect, we recomputed the TAR on the resting recording (Figure 2B). Contrary to that concern, the resting TAR yielded *larger* group differences: means were 0.26 ± 0.19 in CN, 1.59 ± 1.32 in AD (6.1-fold vs CN), and 0.86 ± 0.93 in FTD (3.3-fold). The SSVEP-related bias was therefore *attenuating* rather than exaggerating: the SSVEP inflated alpha preferentially in CN, reducing the apparent group contrast during photic recordings. Both formulations converge on posterior slowing in dementia, with the resting formulation being more sensitive.

**Figure 2:**
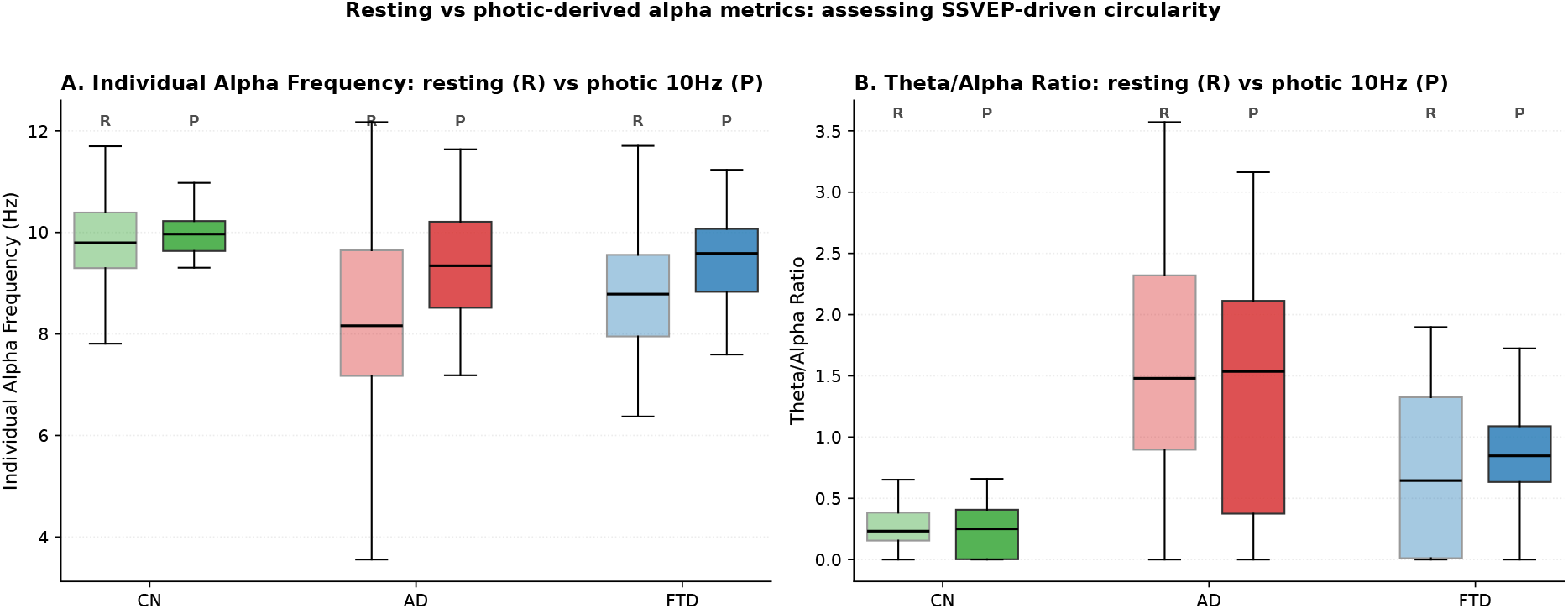
Resting vs photic-derived alpha metrics. (A) Individual alpha frequency computed on the resting recording (R) versus on the photic 10 Hz block (P), revealing that the photic estimate underestimates alpha slowing in dementia. (B) Theta/alpha ratio in resting vs photic conditions; the resting formulation yields larger group effects, confirming that the SSVEP-driven contamination during photic recording is conservative in direction.

### 3.4 Robust alpha slowing on the resting recording

Individual alpha frequency computed on the resting recording (uncontaminated by the SSVEP peak that pulls IAF toward 10 Hz in preserved-driving participants) showed a robust reduction in AD and a smaller, non-significant reduction in FTD. CN averaged 9.82±0.97 Hz, AD averaged 8.30 ± 1.60 Hz (adjusted Δ = −1.54 Hz vs CN, 95% CI [−2.31, −0.77], *p <* 0.001), and FTD averaged 9.14 ± 1.64 Hz (Δ = −0.69 Hz, 95% CI [−1.54, +0.16], *p* = 0.11). These effects are markedly stronger than the photic-derived estimates (9.62±1.01 for AD and 9.55±1.16 for FTD, differences of only ∼ 0.4 Hz; Figure 2A), illustrating that the photic recording underestimates alpha slowing by forcing the spectral peak toward the stimulation frequency in preserved-driving participants. Variance was substantially increased in both dementia groups (Levene’s *p <* 0.001), consistent with the loss of a stable dominant alpha peak in a subset of patients.

### 3.5 Functional connectivity at the photic frequency

wPLI in [9, 11] Hz did not differ between groups in any ROI combination (Figure 1D). Fronto-posterior wPLI was 0.775 ± 0.203 in CN, 0.718 ± 0.166 in AD, and 0.741 ± 0.137 in FTD (two-sided AD-CN *t* = −1.05, *p* = 0.30; FTD-CN *t* = −0.53, *p* = 0.60; AD-FTD *t* = −0.43, *p* = 0.67; Cohen’s *d* ranging −0.15 to −0.31). Similar null results were obtained for posterior intra-ROI (0.762, 0.737, 0.747; both *p >* 0.27), centro-central (0.735, 0.725, 0.691; both *p >* 0.56), and fronto-frontal (0.744, 0.711, 0.749; both *p >* 0.43). wPLI retained 22 CN, 24 AD, and 13 FTD subjects with sufficient epochs.

Because absence of a significant difference is not evidence of equivalence, we conducted TOST equivalence testing on the fronto-posterior contrast using the same ±0.5 SD margin adopted for the primary AD-versus-FTD DI equivalence analysis. TOST failed to establish equivalence in any pairwise comparison (AD vs CN *p* = 0.26; FTD vs CN *p* = 0.19; AD vs FTD *p* = 0.16); the 90% confidence intervals for the group differences all crossed the equivalence bounds. The wPLI analysis is therefore *underpotrered* to distinguish between genuine preservation and small-to-moderate group differences, particularly for FTD (*n* = 13). A further caveat is that wPLI was computed on the same ∼5 s epochs whose within-subject reliability was near zero for the DI at 10 Hz (Section 3.2); the estimational variance of wPLI in short epochs is known to be substantial[50], so per-subject wPLI values likely inherit similarly limited reliability, further attenuating the study’s ability to detect group-level differences.

Taken together, the wPLI results should be read as “no group difference detected under the current protocol, with insufficient evidence to distinguish preservation from small effects,” rather than as positive evidence for preserved phase coupling.

### 3.6 Sex-related moderation

In males, the DI reduction at 10 Hz was numerically larger (AD-CN *β* = −9.35; FTD-CN *β* = −7.34) but non-significant due to reduced subgroup size (*n* = 38, both *p >* 0.13). In females the DI reduction was attenuated (AD-CN *β* = −0.63, *p* = 0.82; FTD-CN *β* = −1.80, *p* = 0.60, *n* = 42). TAR remained significant in both sexes (males AD-CN *p* = 0.034; females AD-CN *p* = 0.025). Three explanations are non-exclusive: females have higher baseline alpha reducing the DI contrast; AD was 69% female generating residual confounding; and subgroup-level sample sizes yield imprecise estimates.

### 3.7 Classification performance and incremental value

LOSO with SVM-RBF (primary model): balanced accuracy was 0.577 [0.472, 0.680] with resting-only, 0.428 [0.328, 0.527] with photic-only, and 0.543 [0.442, 0.639] combined (Table 3; Figure 3). Bootstrap CIs excluded chance-level (1/3) for every model with resting-state information. Permutation test on SVM-RBF combined (500 permutations) yielded a null distribution centered at 0.326 (SD 0.069) and a one-sided *p* = 0.002 (Figure 5), confirming above-chance discrimination.

**Table 3:** LOSO classification performance. Balanced accuracy with 95% bootstrap CI (10,000 iterations). SVM-RBF declared primary model; XGBoost AUC of 0.33 falls below the chance-level of 0.5 indicating inverted probability calibration. Permutation test on SVM-RBF combined: *p* = 0.002. McNemar test for incremental value (combined vs resting-only, SVM-RBF): *p* = 0.85.

| Feature set | Model | Balanced Acc. | 95% CI |
| --- | --- | --- | --- |
| Resting only | LogReg Elastic Net | 0.500 | [0.394, 0.605] |
|  | **SVM-RBF** | **0.577** | **[0.472, 0.680]** |
|  | Random Forest | 0.510 | [0.408, 0.616] |
|  | Gradient Boosting | 0.389 | [0.297, 0.484] |
|  | XGBoost | 0.417 | [0.320, 0.518] |
| Photic only | LogReg Elastic Net | 0.369 | [0.274, 0.469] |
|  | SVM-RBF | 0.428 | [0.328, 0.527] |
|  | Random Forest | 0.422 | [0.339, 0.501] |
|  | Gradient Boosting | 0.408 | [0.314, 0.502] |
|  | XGBoost | 0.406 | [0.312, 0.501] |
| Combined | LogReg Elastic Net | 0.451 | [0.351, 0.555] |
|  | **SVM-RBF** | **0.543** | **[0.442, 0.639]** |
|  | Random Forest | 0.501 | [0.419, 0.580] |
|  | Gradient Boosting | 0.535 | [0.447, 0.627] |
|  | XGBoost | 0.582 | [0.493, 0.672] |

**Figure 3:**
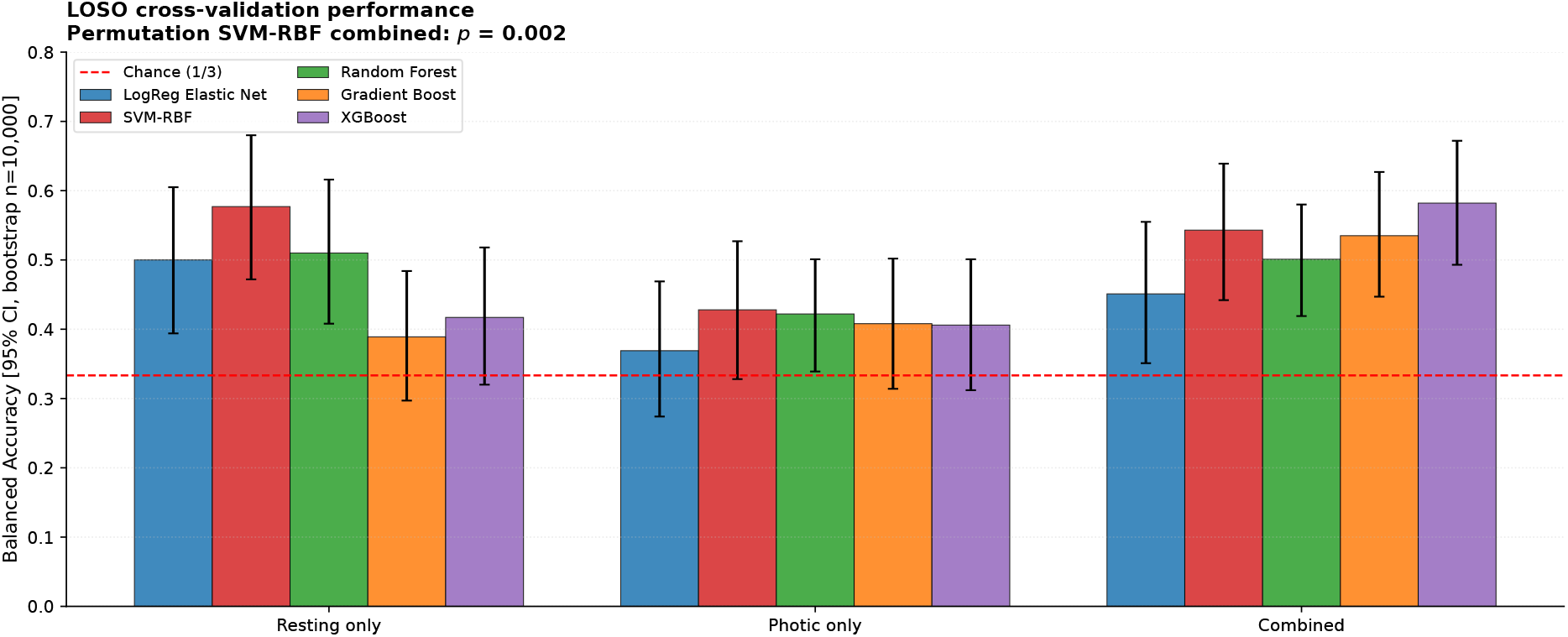
LOSO classification performance. Balanced accuracy with 95% bootstrap confidence intervals for five models across three feature sets. Combined-model performance does not significantly exceed resting-only (McNemar *p* = 0.85).

**Figure 4:**
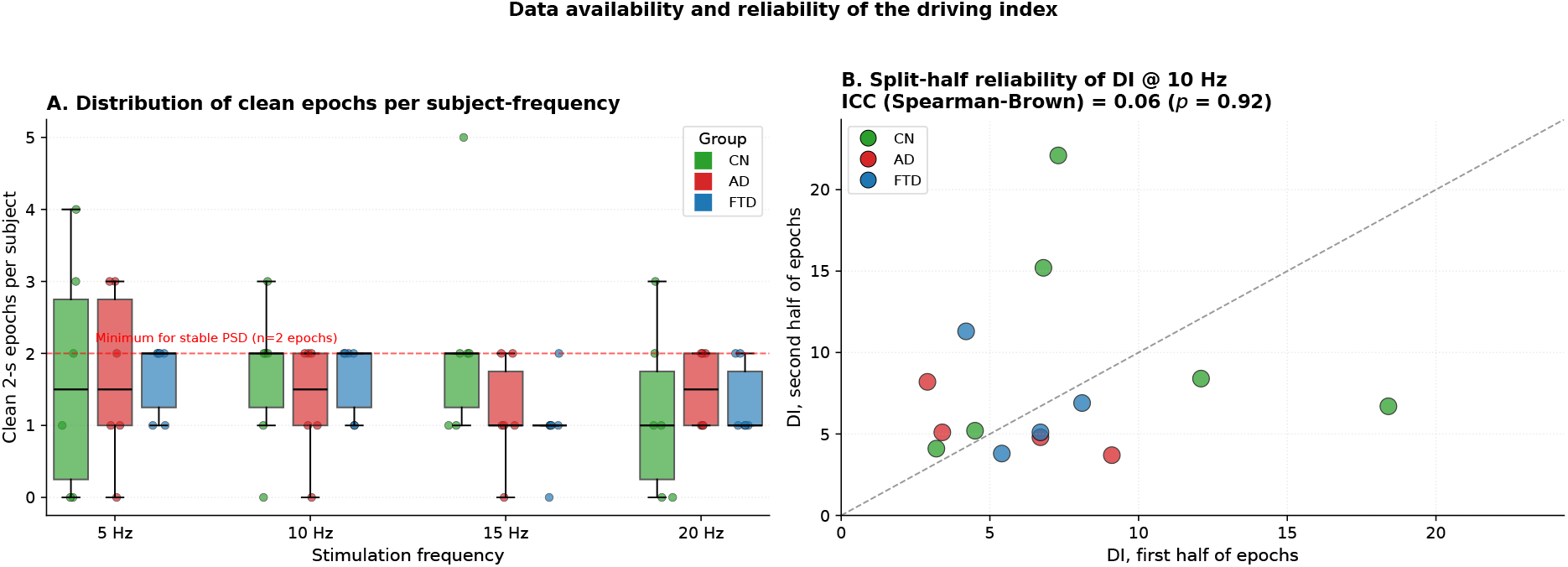
Data availability and reliability ofthe driving index. (A) Distribution of clean 2-second epochs per subject and per stimulation frequency (mean 1-3 epochs per cell). (B) Split-half reliability scatter for the DI at 10 Hz (n = 11 subjects with ≥ 2 epochs per half): Spearman-Brown corrected ICC = 0.06 indicates essentially null within-subject reliability at the individual level.

**Figure 5:**
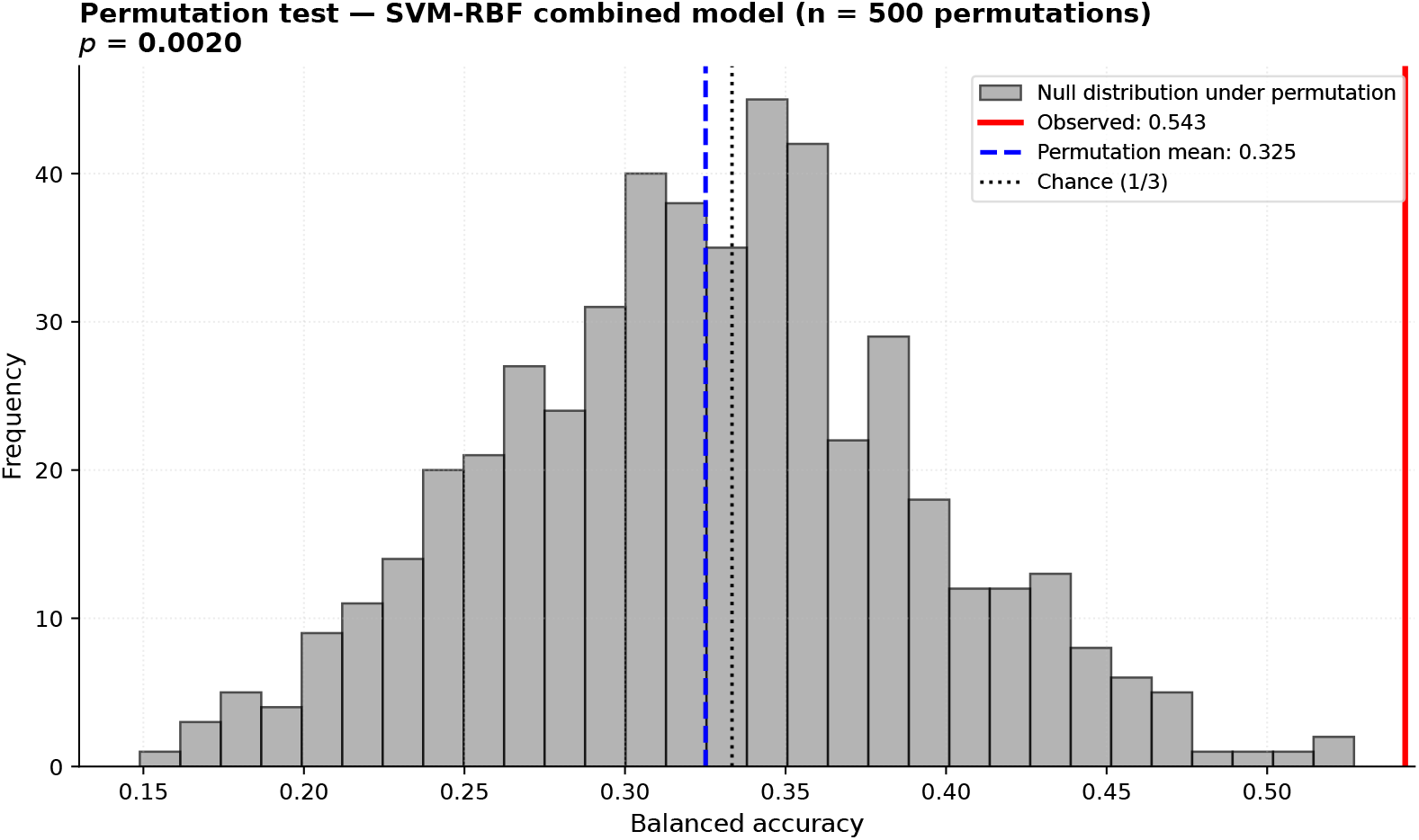
Permutation test. Null distribution of balanced accuracy for the SVM-RBF combined model under 500 label permutations. Observed accuracy (red) significantly exceeds the null distribution (*p* = 0.002).

The pre-specified McNemar exact test comparing SVM-RBF resting-only vs combined found no significant improvement (*p* = 0.85): of 83 subjects, 36 were correctly classified by both, 21 misclassified by both, 12 correctly classified only by combined, and 14 correctly classified only by resting-only. Photic features did not provide incremental classification value.

### 3.8 Within-AD severity

Spearman correlation between photic 10 Hz DI and MMSE within AD (*n* = 28) was small and non-significant (*ρ* = 0.13, *p* = 0.53). Multiple regression of DI on MMSE adjusting for age yielded a similar conclusion (*β*_MMSE_ = +0.085, *p* = 0.57). Within-AD analyses for TAR and IAF were likewise null. The photic driving deficit within AD operates as a threshold marker rather than a dose-response indicator of severity.

## 4 Discussion

### 4.1 Summary of principal findings

Three findings emerge with mutually consistent direction. First, the photic driving index at 10 Hz is reduced approximately 50% in both AD and FTD relative to CN (*β*_AD×10Hz_ = −6.96, *p* = 0.004; *β*_FTD×10Hz_ = −6.74, *p* = 0.012), and this reduction remained significant under log-transformation and 5/95% winsorization, indicating that the effect is not driven by a small number of extreme values. Responses to 5, 15, and 20 Hz do not differ from controls, defining a selective alpha-band deficit rather than a general loss of cortical reactivity. Second, resting-state analyses reveal robust posterior slowing—IAF reduced by 1.5 Hz in AD and posterior TAR elevation reaching 6.1-fold in AD (3.3-fold in FTD)—confirming that the SSVEP-derived estimates conservatively underestimated the underlying phenomenon. Third, despite strong mechanistic signals, the combined photic-plus-resting feature set did not improve LOSO classification beyond resting alone (McNemar *p* = 0.85), and split-half reliability of the DI at the individual level was essentially null (ICC = 0.06, though estimated on only *n* = 11 and hence with wide uncertainty) . Together these describe a *mechanistic biomarker with detectable population-level effects but limited subject-level reliability under the clinical photic protocol*.

### 4.2 A selective alpha-band deficit shared between AD and FTD

The selectivity of the deficit to the 10 Hz frequency is the most novel feature of our results. Classical quantitative studies reported reductions in AD alpha-band responsiveness using coherence-based methods[27-29, 31]; multiscale-entropy analyses confirmed reduced adaptability to repetitive input[33, 34]. Our mixed-effects formulation extends this literature by demonstrating that the deficit is frequency-tuned: the same patients who fail to amplify 10 Hz input respond comparably to controls at 5, 15, and 20 Hz. By including FTD in the same model we show the frequency-tuned attenuation is equally present in FTD, a population that classical visual-EEG literature characterized as “relatively preserved”[35] and that had received little quantitative investigation in this framework.

### 4.3 The AD ≈ FTD phenotype: shared vulnerability with divergent neuropathology

The most clinically counterintuitive finding is the indistinguishable magnitude of the deficit between AD and FTD (Hedges’ *g* = −0.02; TOST-confirmed equivalence within ±4 DI units, *p* = 0.027). Three non-exclusive mechanisms could converge on this shared posterior phenotype. First, a common cholinergic contribution is unlikely to fully account for it. Short-latency afferent inhibition (an in-vivo cholinergic marker) is *reduced in AD but preserved in FTD*[57], indicating basal-forebrain cholinergic loss is not a shared feature. This weakens the simplest mechanistic narrative.

Second, network-level convergence is a more plausible candidate. Multimodal connectivity studies show that AD and FTD, despite divergent proteinopathies, affect partially overlapping posterior default-mode network nodes[58, 59]. Graph-theoretical analyses of resting EEG indicate loss of small-world properties in AD[60], with recent extensions revealing convergent and divergent network signatures across AD and FTD[23, 24].

Third, selective vulnerability of thalamic alpha pacemakers is consistent with the dissociation between reduced amplitude and preserved phase coupling: pulvinar and reticular thalamic neurons pacing alpha[26] could undergo gain reduction without losing synchronizing influence.

### 4.4 Amplitude reduction and the mechanistic role of phase coupling: a hypothesis under limited evidence

The dissociation between the robust group-level DI reduction and the absence of detectable group differences in wPLI is mechanistically *suggestive* but does not, on the present evidence, establish a mechanism. Two qualitatively distinct network dysfunctions can produce a reduced SSVEP: loss of cortical *gain* (cortex receives thalamic drive but amplifies less) or loss of network phase synchrony. Our wPLI results are compatible with the first account, but critically cannot rule out the second, because TOST equivalence testing did not establish preservation and the reliability of wPLI over short epochs is likely limited (Section 3.5). Biophysical models of alpha rhythm generation in AD propose that a dominant pathological change is a reduction in gain rather than a disorganization of network coupling[61], and our data are consistent with that model without confirming it. We therefore frame the “gain reduction over preserved coupling” account as a mechanistic hypothesis to be tested prospectively with longer-duration photic recordings and equivalence-powered connectivity analyses. If this hypothesis is corroborated, the clinical implication is that gain-restoration interventions—sensory entrainment[41–43], pharmacological modulation of NMDA-receptor cortical excitability—might restore driving without requiring restoration of coupling. If it is not, the same interventions may still be relevant but the mechanistic rationale would shift accordingly.

### 4.5 Theta/alpha ratio: convergent evidence with methodological refinement

Consistent with external methodological review, we recomputed both TAR and IAF on the resting recording. The resting-based estimates yielded *stronger* group effects than photic-based estimates: resting TAR showed 6.1-fold elevation in AD (vs 3.8-fold in photic), and resting IAF showed 1.5 Hz reduction in AD (vs 0.4 Hz in photic). The direction of SSVEP-driven contamination was therefore *conservative*: the photic recording underestimated posterior slowing because the SSVEP artificially elevated alpha power in preserved-driving controls. Reporting both formulations strengthens interpretation of the two metrics as genuinely convergent indicators of posterior alpha dysfunction and clarifies that driving-index and slowing findings are conceptually related but computationally distinct.

### 4.6 Mechanism without incremental diagnostic value: reliability as the missing link

In sharp contrast to the strong mechanistic signal, photic features did not improve classification beyond resting EEG (McNemar *p* = 0.85). This dissociation is now mechanistically understood: DI has moderate reliability at 5 Hz (ICC_SB_ = 0.66) but essentially null reliability at 10 Hz (ICC_SB_ = 0.06) in the clinical photic protocol, because each subject provides only 1-3 clean 2-s epochs per frequency. This within-subject unreliability coexists with a robust across-subject effect detectable only through mixed-effects modeling that borrows strength across subjects. The mechanism revealed by our analyses is a real group-level neurophysiological phenomenon—posterior alpha-circuit dysfunction shared between AD and FTD—but it is not, in the current protocol, a measurable biomarker at the individual level. Photic protocols with substantially longer eyes-closed segments per frequency (30–60 s) might reach subject-level ICC compatible with individual inference; this is a testable design recommendation.

### 4.7 Methodological lessons for EEG machine learning on open data

Recent machine-learning studies on the ds004504 dataset have reported classification accuracies of 95–99%[23, 24, 36, 37, 40]. Our subject-level LOSO balanced accuracy of∼ 0.58 falls well below these figures. The discrepancy is fully explained by validation strategy: when cross-validation is performed at epoch level, the model learns subject-specific spectral fingerprints producing inflated estimates that do not generalize[46]. Our subject-level LOSO with 10,000-fold bootstrap CIs provides a methodologically transparent baseline for future work.

The XGBoost AUC anomaly deserves comment: with combined features it yielded balanced accuracy 0.582 but AUC 0.333, *below* the invariant chance level of 0.5 for a probabilistic AUC[55], evidencing systematically inverted probability calibration rather than random-guess behavior. This would have been undetected with a single metric; reporting balanced accuracy, macro-F1, and AUC together makes calibration failure visible.

### 4.8 Sex-related moderation

Three non-exclusive explanations for the sex moderation of the photic effect. *Biological*: females exhibit higher baseline alpha amplitude[22], mathematically reducing the DI contrast at the stimulus frequency. TAR, as a power-band ratio, is less sensitive. *Methodological*: AD was 69% female generating residual confounding despite adjustment. *Statistical*: subgroup sizes (*n* ≈ 5-12 per cell) yield imprecise estimates. Sex moderation should be characterized in deliberately balanced cohorts before any sex-specific biomarker is proposed.

### 4.9 Translational perspective: from passive probe to active therapy

While our clinical protocol caps at 20 Hz precluding gamma-band examination, the broader photic-stimulation-in-dementia landscape has shifted from diagnostic tool to candidate therapy. The seminal demonstration that 40 Hz stimulation reduces amyloid load and microglial dysregulation in murine AD models[41], with extensions showing multimodal benefit[42] and gamma-mediated neuroprotection[43], has opened a new translational axis. Recent advances in comfortable 40 Hz delivery[44, 45] reduce practical barriers to chronic deployment. Our finding of impaired alpha-band driving with preserved phase-coupling scaffolding suggests that posterior alpha and gamma circuits may be differentially vulnerable, and that phase-coupling integrity is a relevant pre-treatment biomarker for gamma-entrainment trials.

### 4.10 Limitations

Several limitations qualify our findings. *Sample*: single-site cohort with clinically rather than biomarker-confirmed diagnoses; sex imbalance may leave residual confounding. *Recording protocol*: short eyes-closed segments (∼ 5 s per frequency) limit per-frequency spectral stability and the protocol caps at 20 Hz. *Spatial resolution*: 19-channel montage precludes source-level reconstruction. *Cross-sectional design*: prevents evaluation of the photic deficit as an early biomarker of progression. *Medication and comorbidity* data are unavailable.

#### Within-subject reliability ofthe driving index

the clinical protocol provides only 1–3 clean 2-s epochs per subject per stimulation frequency, resulting in null split-half reliability at 10 Hz (ICC = 0.06). Group-level effects are nevertheless detectable through mixed-effects modeling; however, deployment as an individual biomarker would require substantially extended per-frequency windows.

### 4.11 Future directions

Three priorities. First, replication in independent cohorts with biomarker-confirmed diagnoses. Second, extension of the photic protocol into the gamma band, simultaneously testing gamma-entrainment circuit integrity (a therapeutic target in AD[41–43]) and enabling direct comparison with alpha-band findings. Third, longitudinal designs to test whether the deficit precedes cognitive impairment. Methodologically, we recommend construction of adversarial-LOSO benchmarks on existing open data, requiring subject-level metrics in pre-registered formats.

### 4.12 Conclusion

Photic stimulation reveals a selective alpha-band driving deficit and elevated theta/alpha ratio shared between AD and FTD, providing mechanistic information about a common posterior thalamocortical vulnerability partially independent of divergent neuropathological substrates. The absence of detectable group differences in phase-lag connectivity is *compatible* with—but does not formally demonstrate—a dysfunction predominantly localized to cortical gain rather than network synchronization; disentangling these accounts is left to prospective work with longer photic recordings adequately powered for equivalence testing on connectivity. Photic features do not provide incremental classification value over resting-state EEG, a limitation now mechanistically explained by the null within-subject reliability of the DI in the clinical protocol. Under the present protocol, therefore, the clinical role of intermittent photic stimulation in dementia is mechanistic rather than diagnostic. These findings refine the clinical narrative around photic stimulation, expose the over-optimism of epoch-level validation on open data, and identify posterior alpha generator integrity as a candidate pre-treatment marker for gamma-entrainment therapeutic trials.

## Acknowledgments

We thank the original investigators at the 2nd Department of Neurology, AHEPA University Hospital, Thessaloniki, for releasing the ds004504 and ds006036 datasets under CC0 license. We thank Universidad CES and the Grupo de Neurociencias de Antioquia (GNA), Universidad de Antioquia, for institutional support.

## Data and code availability

The datasets analyzed are publicly available under the CC0 1.0 license on OpenNeuro at https://openneuro.org/datasets/ds004504. and https://openneuro.org/datasets/ds006036. All analysis code, derived subject-level feature matrices, statistical outputs, sensitivity and equivalence analyses, and publication figures are openly available at https://github.com/pedrovelezpardo/EEG_Photic_AD_FTD released with tag `v1. 0-submission`. The repository includes a `CITATION`. cff file for citation and an MIT license for the analysis code.

## Author contributions

The following are the CRediT (Contributor Roles Taxonomy) contributions of each author. **Pedro Velez-Pardo:** Conceptualization, Data curation, Formal analysis, Investigation, Methodology, Software, Validation, Visualization, Project administration, Writing – original draft, Writing - review and editing.

**Ricardo Montoya Monsalve:** Conceptualization, Investigation, Writing – review and editing. **Daniel Vasquez:** Formal analysis, Methodology, Validation, Writing – review and editing. **Sofía Campuzano Cortina:** Conceptualization, Investigation, Writing – review and editing. **Eduardo Montoya Guevara:** Methodology, Supervision, Validation, Writing – review and editing.

All authors have read and agreed to the submitted version of the manuscript.

## Declaration of generative AI and AI-assisted technologies in the manuscript preparation process

During the preparation of this work, the authors used Claude (Anthropic, PBC, San Francisco, CA, USA, https://claude.ai, accessed July 2026) in order to (i) review, refactor, and debug portions of the analysis code (Python and R), (ii) assist with drafting, iterative revision, and language polishing of the manuscript and companion submission documents (cover letter, highlights, README), and (iii) reformat the manuscript to the Biomedical Signal Processing and Control author guidelines. After using this tool, the authors reviewed and edited the content as needed and take full responsibility for the content of the published article.

