## Supplementary material for "Selective 10-Hz photic driving deficit shared between Alzheimer’s and frontotemporal dementia"

Universidad CES and Grupo de Neurociencias de Antioquia (GNA), Universidad de Antioquia, Medellín, Colombia.

This document contains the supplementary tables (S1–S4) referenced in the main manuscript. Raw data sources: OpenNeuro ds004504 and ds006036 (CC0). Analysis code and derived features: https://github.com/pedrovelezpardo/EEG_Photic_AD_FTD (tag v1.0-submission).

### Table S1. Distribution of clean 2-s epochs per subject and stimulation frequency.

**Table S1:** *Summary statistics of the number of clean 2-s epochs available per subject at each stimulation frequency, in a representative subsample of 18 subjects (6 per diagnostic group) used for the split-half reliability estimation. Values are mean, SD, minimum and maximum per subject.*

| Group | Freq (Hz) | Mean | SD | Min | Max | n |
|---|---|---|---|---|---|---|
| AD | 5 | 1.67 | 1.21 | 0 | 3 | 6 |
| AD | 10 | 1.33 | 0.82 | 0 | 2 | 6 |
| AD | 15 | 1.17 | 0.75 | 0 | 2 | 6 |
| AD | 20 | 1.50 | 0.55 | 1 | 2 | 6 |
| CN | 5 | 1.67 | 1.63 | 0 | 4 | 6 |
| CN | 10 | 1.67 | 1.03 | 0 | 3 | 6 |
| CN | 15 | 2.17 | 1.47 | 1 | 5 | 6 |
| CN | 20 | 1.17 | 1.17 | 0 | 3 | 6 |
| FTD | 5 | 1.67 | 0.52 | 1 | 2 | 6 |
| FTD | 10 | 1.67 | 0.52 | 1 | 2 | 6 |
| FTD | 15 | 1.00 | 0.63 | 0 | 2 | 6 |
| FTD | 20 | 1.33 | 0.52 | 1 | 2 | 6 |

### Table S2. Split-half reliability of the driving index by stimulation frequency.

**Table S2:** *Split-half reliability (Spearman-Brown corrected ICC and Pearson correlation) of the driving index at each stimulation frequency, computed on the subsample of subjects with at least two clean epochs available per half.*

| Frequency (Hz) | n subjects | Pearson r | ICC (SB) | p-value |
|---|---|---|---|---|
| 5 | 10 | 0.495 | 0.662 | 0.146 |
| 10 | 11 | 0.033 | 0.064 | 0.923 |
| 15 | 7 | -0.249 | -0.662 | 0.591 |
| 20 | 7 | 0.207 | 0.343 | 0.656 |

### Table S3A. Sensitivity analyses of the group effect on the driving index at 10 Hz.

**Table S3A:** *Mixed-effects marginal contrast at 10 Hz (AD vs CN and FTD vs CN) under three specifications: raw DI, log(1+DI), and 5/95% winsorization per stimulation frequency. All contrasts adjusted for age, sex, and number of clean epochs, with random intercept by subject.*

| Analysis | Transformation | β_AD | SE | p | 95% CI | β_FTD | SE | p | 95% CI |
|---|---|---|---|---|---|---|---|---|---|
| Primary | Raw DI | -6.24 | 2.16 | 0.004 | [-10.47, -2.00] | -6.16 | 2.36 | 0.009 | [-10.78, -1.54] |
| Sensitivity 1 | log(1+DI) | -0.53 | 0.21 | 0.010 | [-0.94, -0.13] | -0.59 | 0.23 | 0.009 | [-1.03, -0.15] |
| Sensitivity 2 | Winsorized 5/95% | -3.40 | 1.55 | 0.028 | [-6.43, -0.37] | -3.46 | 1.69 | 0.041 | [-6.76, -0.15] |

### Table S3B. Rank-based sensitivity — Kruskal-Wallis and post-hoc pairwise Mann-Whitney tests.

**Table S3B:** *Kruskal-Wallis omnibus test on the raw DI at 10 Hz, with pairwise Mann-Whitney post-hoc comparisons and Bonferroni-corrected p-values across three comparisons.*

| Test | Statistic | p | p (Bonferroni) | Interpretation |
|---|---|---|---|---|
| Kruskal-Wallis (global) | H = 7.44 | 0.024 | — | Significant global group effect |
| Mann-Whitney CN vs AD | U = 546 | 0.022 | 0.067 | Marginal |
| Mann-Whitney CN vs FTD | U = 367 | 0.018 | 0.055 | Marginal |
| Mann-Whitney AD vs FTD | U = 335 | 0.636 | 1.000 | No difference (consistent with equivalence) |

### Table S4. TOST equivalence testing for wPLI at 10 Hz (fronto-posterior).

**Table S4:** *Two One-Sided Tests (Schuirmann/Lakens) for pairwise equivalence of the fronto-posterior weighted phase-lag index at 10 Hz, with a Cohen's d = 0.5 equivalence margin. TOST p ≥ 0.05 indicates that formal equivalence is not established.* $n_1$ *and* $n_2$ *correspond to the number of subjects retained in each group with sufficient epochs for wPLI computation.*

| Comparison | $n_1$ | $n_2$ | Δ mean | Cohen's d | 90% CI | Bound (d=0.5) | TOST p | Interpretation |
|---|---|---|---|---|---|---|---|---|
| AD vs CN | 24 | 22 | -0.057 | -0.309 | [-0.149, +0.035] | ±0.092 | 0.260 | No equivalence established |
| FTD vs CN | 13 | 22 | -0.034 | -0.186 | [-0.142, +0.074] | ±0.091 | 0.188 | No equivalence established |
| AD vs FTD | 24 | 13 | -0.023 | -0.147 | [-0.114, +0.068] | ±0.078 | 0.156 | No equivalence established |

**Note:** *Abbreviations: AD, Alzheimer's disease; FTD, frontotemporal dementia; CN, cognitively normal controls; DI, driving index; TOST, two one-sided tests; ICC (SB), Spearman-Brown corrected intraclass correlation coefficient; wPLI, weighted phase-lag index.*
